# Factors associated with COVID-19 vaccine hesitancy in Senegal: a mixed study

**DOI:** 10.1101/2021.12.20.21267724

**Authors:** Mouhamadou Faly Ba, Adama Faye, Babacar Kane, Amadou Ibra Diallo, Amandine Junot, Ibrahima Gaye, Emmanuel Bonnet, Valéry Ridde

**Affiliations:** Institute of Health and Development (ISED), Cheikh Anta Diop University, Dakar, Senegal; STAPS Department, Faculty of Human and Environmental Sciences, La Réunion University, 97430 Le-Tampon, Réunion; IRD, UMR 215 Prodig, 5, cours des Humanités, Aubervilliers, France; CEPED, IRD-University of Paris, ERL INSERM SAGESUD, Paris, France

**Author notes:** Corresponding author: (MF Ba).

**Keywords:** Vaccine hesitancy, COVID-19, Mixed method, Senegal

## Abstract

**Introduction:** The most effective way to control the COVID-19 pandemic in the long term is through vaccination. Two of the important components that can hinder it are vaccine hesitancy and vaccine refusal. This study, conducted before the arrival of the vaccines in Senegal, aims to assess and identify factors associated with hesitancy to the COVID-19 vaccine.

**Methods:** This study was an explanatory, sequential, mixed-methods design. We collected quantitative data from December 24, 2020, to January 16, 2021, and qualitative data from February 19 to March 30, 2021. We conducted a marginal quota sampling nationwide. We used a structured questionnaire to collect data for the quantitative phase and an interview guide with a telephone interview for the qualitative phase. We performed descriptive, bivariate, and multivariate analyses with R software version 4.0.5 for the quantitative phase; and performed manual content analyses for the qualitative phase.

**Results:** We surveyed 607 people for the quantitative phase, and interviewed 30 people for the qualitative phase. Individuals who hesitated or refused to be vaccinated represented 12.9% and 32.8%, respectively. Vaccine hesitancy was related to gender, living in large cities, having a poor attitude towards the vaccine, thinking that the vaccine would not help protect them from the virus, being influenced by people important to them, and lacking information from health professionals. Vaccine refusal was related to living in large cities, having a poor attitude towards the vaccine, thinking that the vaccine would not help protect them from the virus, thinking that the vaccine could endanger their health, trusting opinions of people who were important to them, and lacking information from health professionals.

**Conclusion:** The results of the study show that the factors associated with hesitancy and refusal to be vaccinated against COVID-19 are diverse and complex. Reducing them will help to ensure better vaccination coverage if the current challenges of vaccine accessibility are addressed. Therefore, governments and health authorities should intensify their efforts to promote vaccine confidence and reduce misinformation.

## Introduction

Coronavirus disease 2019 (COVID-19) remains a significant public health concern. Although much effort has been devoted to implementing control strategies—including travel bans, isolation of confirmed cases and contacts, social distancing, and hygiene measures—virus transmission is likely to rebound when these strategies are lifted [1]. Among multiple possible strategies, one way to control this pandemic is through mass vaccination [2]. Achieving effective results from vaccination depends not only on accessibility, which remains a major challenge in Africa, but also on the acceptance and willingness of the population to be vaccinated. [3]. Thus, one of the major obstacles to achieving high immunization coverage is vaccine hesitancy [4]. Beyond the long-standing debates on the concept and its scope [5], the World Health Organization (WHO) defines vaccine hesitancy as the delay in acceptance or refusal of vaccination despite the availability of immunization services [6].

Worldwide, studies show very high variability in acceptability of the COVID-19 vaccine, with rates ranging from 29.4% to 86.0%. [7–13]. In the majority of studies of the public stratified by country, acceptance of COVID-19 vaccination showed a level ≥70% [14]. A survey of 15 African countries showed that approximately 80% of people are willing to accept the COVID-19 vaccine once it is available and is considered safe and effective. Although the overall results are encouraging, there are significant regional differences in Africa [15]. A meta-analysis showed that the proportion of individuals reporting that they would refuse a COVID-19 vaccine was 14.3% [95% CI: 11.4% to 17.9%], and the proportion reporting uncertainty was 22.1% [95% CI: 17.8% to 27.1%]. [16]. The latter also showed that intentions to vaccinate have decreased over time while refusals have increased [16]. Several factors can influence acceptance or refusal of the vaccine (professional status, politics, gender, age, education, income, etc.). [17]. In addition, the novelty of the disease, concerns about the safety and efficacy of the vaccine, and distrust of governments have resulted in a significant proportion of people indicating a reluctance to be vaccinated against COVID-19 [17].

Senegal launched its vaccination campaign against COVID-19 on 23 February 2021. As of December 2, 2021, 1,328,633 people have received at least one dose, including 924,182 people who have received two doses, representing a complete coverage of 5.5% of the total population. [18]. This coverage is far from the objective set by the authorities, which was to ensure vaccination of at least 20% of the general population before June 2021 [19]. One of the important components of this challenge, despite its multifaceted nature, is vaccine hesitancy and refusal. Thus, assessing its scope and magnitude is necessary to guide interventions to build and sustain responses to this epidemic. Understanding and responding to the determinants is necessary to achieve a high vaccine coverage.

This study aims to assess and identify factors associated with hesitancy towards the COVID-19 vaccine in Senegal.

## METHODOLOGY

### Research specifications, data collection and study population

This study is a sequential, explanatory, mixed-methods design where qualitative data should help understand the results of the analysis of the previously collected quantitative data [20]. The writing of the article followed the quality criteria proposed by the Mixed Methods Appraisal Tool [21]. The quantitative data were collected from December 24, 2020, to January 16, 2021, before the vaccine campaign; and the qualitative data were collected during the vaccine campaign - from February 19 to March 30, 2021.

### Sampling

The study population consisted of individuals from the general population living in Senegal aged 18 years and older with a mobile phone number. In June 2020, we conducted an initial nationwide telephone survey of 813 people to measure the social acceptability of governmental measures to control COVID-19 [22]. The study used a marginal quota sampling strategy [23]. In order to have a representative sample of the population, we carried out a stratification according to the weight of the population by region, gender, and age group. Based on this first survey, which did not concern the vaccination aspects, we organized a second survey among these same people. The final quantitative sample size was 607 (74.6%). A comparison of the characteristics of the quotas chosen to constitute the sample between the two surveys shows that while they are not statistically different for region (p=0.99) or age (p=0.08), they are for gender (p=0.04).

The qualitative sample was composed of 30 individuals selected from those who said they were reluctant (n=15) or unwilling (n=15) to be vaccinated, nested within the final quantitative sample (n=607) (Appendix 1). The selection of these 30 individuals followed the same stratification as the quantitative sample to have diverse views. These individuals were drawn at random from the quantitative sample and according to this stratification. Individuals were replaced when they refused to participate or could not be reached.

### Data collection

The quantitative data collection tool was a structured and closed questionnaire. Five female interviewers speaking six languages (French, Diola, Wolof, Sérére, Pulaar, Soninké) collected the data. The survey was conducted by telephone. The interviewers used tablets equipped with an Open Data Kit (ODK) software to administer the questionnaire [24]. We performed data quality control by training interviewers, pre-testing the tools, scanning the questionnaire, collecting the data on a tablet, and recruiting a supervisor to monitor data collection in real-time daily.

Intention to be vaccinated, the dependent variable, was measured with a 5-point Likert scale (strongly agree = 5 to strongly disagree = 1). Following the WHO definition, it was transformed into a 3-mode variable:

- Strongly agree and agree = intention to be vaccinated
- Neither agree nor disagree = reluctant to get vaccinated
- Disagree and strongly disagree = refusal to be vaccinated

The independent variables collected in the quantitative survey were conceptualized according to the WHO vaccine hesitancy model [6]. It concerned:

- Contextual factors: age, gender, region, wealth quintile, education, belief in the safety of the pharmaceutical industry, belief in the accessibility of health personnel to get vaccinated, agreement that there is something wrong with the vaccines, and total trust in the government to fight the epidemic.
- Individual and group influences: perceived importance of getting vaccinated, perceived usefulness of getting vaccinated, perceived responsibility of getting vaccinated, perceived safety of future vaccine, perceived desirability of getting vaccinated, perceived benefits and risks of the vaccine, social influence for receiving the vaccine, trust in health care providers for receiving the vaccine, getting regular information about the vaccine in the coming months, and perceived need for health care workers to provide information about the vaccine.
- Vaccine-specific factors: free vaccines for the entire population

The independent variables composed in the form of a 5-point Likert scale were transformed into binary variables (Yes = Strongly agree and agree; No = Other). For the variable “Confidence in the government in the fight against COVID-19,” which was in the form of a score ranging from 0 to 10, the person was considered to have had complete confidence when he/she had the maximum score. Using principal component analysis (PCA), we obtained the wealth quintile on durable asset ownership and housing characteristics. This approach ranked individuals from the poorest (1) to the least poor (5) to capture the socio-economic differences.

All this made it possible to determine the level of refusal and reluctance to be vaccinated and to identify the factors associated with them.

The qualitative survey was guided by the results of the quantitative analyses by seeking to understand more deeply the reasons for hesitation or refusal. Using an open-ended guide, individual interviews were conducted over the telephone for an average of 30 minutes.

### Data analysis

We performed quantitative analyses with R software version 4.0.5. Categorical variables were described by numbers and percentages. We used the Chi2 test to compare proportions with a 5% alpha risk. We modelled vaccine hesitancy and refusal using multinomial logistic regression in the multivariate analysis. We included all variables with p-values less than 0.25 in the bivariate analysis in the initial model [25]. To construct the final model, we used the stepwise top-down selection procedure in each model. We individually removed variables that did not improve the model [26]. We used the likelihood ratio test to compare the nested models [26]. We used this multivariate analysis to determine adjusted Odds Ratios and estimated the corresponding 95% confidence intervals (CIs) for all variables.

For the qualitative data, we transcribed the interviews in full in French. Then, we performed a manual content analysis [27]. According to the mixed methods approach, divergences and convergences are highlighted in the presentation of the results [28]. Explanatory elements for vaccine refusal or hesitation that were not considered in the quantitative survey emerged in the qualitative survey.

## Results

### Descriptive analysis

In the study, 67.1% of the individuals were between 25 and 59 years of age. Males accounted for 60.3%. The proportion of respondents with no education was 41.7% (Appendix 2).

Individuals who hesitated and refused to be vaccinated were 12.9% and 32.8%, respectively (Appendix 2). In the qualitative survey, although the majority of respondents remained in their positions: *“I am still in my position because I do not know the components of the vaccine, I know absolutely nothing about the vaccine*” (Female, refusal); others changed their positions due to the emerging situation of a new outbreak of COVID-19 cases during this period: “*I am for the vaccine because it can reduce the severity of the disease and the vaccine is important for the second wave*” (Female, refusal).

### Bivariate analysis

The proportion of vaccine-hesitant among those who thought it was not useful to be vaccinated was 23.3%. This proportion was 9.0% among those who thought it was useful (p<0.001). The proportion of individuals who refused to be vaccinated because the vaccine could endanger their health was higher than the proportion who said the vaccine would not endanger their health (67.9% vs 22.8%, p<0.001) (Appendix 3).

### Multivariate analysis

#### Socio-demographic characteristics

Females were more likely to be reluctant to be vaccinated than males (ORa = 2.49 [95% CI: 1.20 - 5.17]). In addition, individuals living in Senegal’s major cities (Dakar, Diourbel, and Thiès) were more likely to hesitate and refuse to be vaccinated than those living elsewhere (ORa = 2.16 [CI95%: 1.04 - 4.48]; ORa = 2.03 [CI95%: 1.04 - 3.96]) (Table 1)

**Table 1:** Results of the multivariate analysis

| Features | Intention (Ref) vs Hesitation |  |  | Intention (Ref) vs Refusal |  |  |
| --- | --- | --- | --- | --- | --- | --- |
|  | ARO <sup>I</sup> | 95% CI <sup>I</sup> | p-value | ARO <sup>I</sup> | 95% CI <sup>I</sup> | p-value |
| **Sex** |  |  |  |  |  |  |
| Male | 1.00 | - |  | 1.00 | - |  |
| Female | **2.49** | **1.20, 5.17** | **0.014** | 1.77 | 0.90, 3.50 | 0.099 |
| **Region** |  |  |  |  |  |  |
| Remainder | 1.00 | - |  | 1.00 | - |  |
| Dakar-Diourbel-Thiès | **2.16** | **1.04, 4.48** | **0.040** | **2.03** | **1.04, 3.96** | **0.038** |
| **I think it's important to get vaccinated** |  |  |  |  |  |  |
| Yes | 1.00 | - |  | 1.00 | - |  |
| No | **2.72** | **1.16, 6.39** | **0.022** | 2.07 | 0.93, 4.61 | 0.075 |
| **I think it is advisable to get vaccinated against COVID-19** |  |  |  |  |  |  |
| Yes | 1.00 | - |  | 1.00 | - |  |
| No | **16.49** | **6.72, 40.59** | **<0.001** | **17.00** | **7.42, 39.00** | **<0.001** |
| **Getting the COVID-19 vaccine will help protect me from the virus** |  |  |  |  |  |  |
| Yes | 1.00 | - |  | 1.00 | - |  |
| No | **20.10** | **8.06, 50.00** | **<0.001** | **19.30** | **8.49, 44.00** | **<0.001** |
| **The coronavirus vaccine could put my health at risk** |  |  |  |  |  |  |
| Yes | 1.00 | - |  | 1.00 | - |  |
| No | 0.56 | 0.20, 1.62 | 0.299 | **0.20** | **0.09, 0.49** | **<0.001** |
| **Coronavirus vaccine may have side effects** |  |  |  |  |  |  |
| Yes | 1.00 | - |  | 1.00 | - |  |
| No | 1.66 | 0.71, 3.90 | 0.199 | 0.67 | 0.32, 1.40 | 0.299 |
| **When the vaccine is offered, most of the people important to me (family, friends) would think that I need to be vaccinated against COVID-19** |  |  |  |  |  |  |
| Yes | 1.00 | - |  | 1.00 | - |  |
| No | **4.97** | **2.34, 10.50** | **<0.001** | **5.61** | **2.83, 11.1** | **<0.001** |
| **I think it will be easy for me to access the health professional to get the coronavirus vaccine if I want it** |  |  |  |  |  |  |
| Yes | 1.00 | - |  | 1.00 | - |  |
| No | **0.33** | **0.13, 0.84** | **0.020** | **0.22** | **0.09, 0.51** | **<0.001** |
| **Health workers should provide appropriate and necessary information and support for immunization** |  |  |  |  |  |  |
| Yes | 1.00 | - |  | 1.00 | - |  |
| No | **22.4** | **1.71, 293** | **0.018** | **26.2** | **2.29, 301** | **0.009** |

<sup>1</sup> AOR = Adjusted Odds Ratio, CI = Confidence Interval, p < 0.05 in bold

#### Attitudes

Individuals who thought it was not important or not desirable to get vaccinated were more reluctant to get vaccinated (OR = 2.72 [CI95%: 1.16 - 6.39]; OR = 16.49 [CI95%: 6.72 - 40.59]) (Table 1). The qualitative research indicates that this could be explained by rumours: “*I hesitate because of the rumours I heard about the vaccine; that’s the first aspect, the second aspect is that I am confused about the time it takes to create it. That is why I am hesitating for the moment and I am waiting for some time to understand how it will manifest itself in the country*” (Male, hesitant).

“*Some people were saying that the vaccine reduces life expectancy and for others, the vaccine was created to lower the birth rate and harm Africans. In any case, people say that and there are also those who say that you could have the virus after being vaccinated, that’s it*. (Female, hesitant)

Individuals who thought it was undesirable to be vaccinated against COVID-19 were more likely to refuse the vaccine (OR = 17.00 [CI95%: 7.42 - 39.00]) (Table 1). Qualitative analyses confirmed this perception. Individuals spoke of concern to reduce the African population with this vaccine: *“I have heard that some people want to harm Africa because they see a very high birth rate in the population. It is also said that those who control the world consider that Africa has many more people because some men marry four women and have 10 children or each woman gives birth to five children. They demand for each couple to have one or two children and if they don’t manage to prohibit the large number of births, they do everything to decrease the size of our population through vaccines*.” (Male, refusal). On another note, one person said that it was not advisable to talk about COVID-19 vaccination in Africa when the epidemic is taking a greater toll in northern countries: “*They should start with them first. If they had done that until they were cured, until they were stable, then they would say, there is still the part of the Africans, I would have understood*” (Male, refusal)

#### Perceived benefits

Individuals who said that getting vaccinated would not help protect them from the virus were more likely to hesitate and refuse to get vaccinated (OR = 20.10 [CI95%: 8.06 - 50.00]; OR = 19.30 [CI95%: 8.49 - 44.00]) (Table 1). Even though the individuals in the qualitative survey think that there could be a benefit, they address the fact that the vaccine only makes it possible to reduce the risk of seriousness once the person is infected by the virus: “*Ah, as we said about vaccination, today if you vaccinate yourself at least, maybe this disease even if it affects you, it won’t be serious. I can’t say that you won’t get Covid-19. Today, no one can guarantee that this disease will not affect you after vaccination*” (Female, refusal).

“*There can be advantages because it’s the last resort, it’s the last solution. When you get the vaccine, even if you have the disease, you can have the strength to fight it and it won’t harm you. Now we have hope*.” (Male, hesitant)

#### Perceived risks

Individuals who thought that the vaccine might endanger their health were more likely to refuse the vaccine (OR = 5.00 [95% CI: 2.04 - 11.11]) (Table 1). Individuals refusing the vaccine cited adverse effects of some vaccines and deaths observed in some countries after the start of the vaccination campaign as explanations: “*You know, when you follow the news closely, there are certain things about which you can have doubts. I saw on France 2, in their 8 o’clock news, that in Norway they started to vaccinate retired people and most of those who were vaccinated died and it was the French television that showed it. There are some things we have doubts about. Yes you know that this information, the television, if it was another television I can not believe it but France TV TF1 or LCI, han? Yes, the side effects, we have to say so because there are side effects, they have to tell us about the side effects*” (Male, refusal). This situation creates doubt in the respondents and leads them to adopt a precautionary principle to observe the possible positive or negative effects of the vaccine before taking the plunge: “*If people who are vaccinated stay 3 to 4 months without anything happening to them or if they don’t have any undesirable effects, I’ll get vaccinated*” (Female, hesitant)

#### Subjective standards

Individuals who thought that most people important to them would not think they should be vaccinated against COVID-19 when the vaccine was offered were more likely to hesitate and refuse to be vaccinated (OR = 4.97 [95% CI: 2.34 - 10.50]; OR = 5.61 [95% CI: 2.83 - 11.10]) (Table 1). However, the qualitative survey showed that for most individuals hesitating and refusing to be vaccinated, the opinion of their family member would not affect their decision in any way: “*No, if I have to be vaccinated, I will do it willingly, but not under the influence of anyone*” (Female, Hesitant). For others, the opinion of people who have expertise in this field can be a determining factor in changing their behaviour: “*For me, only doctors can influence us because they know it better than us. But for the others, even if they have knowledge, for me, maybe what they say is true, but I trust the doctors more, they know the job better*. (Female, refusal).

#### Information and conspiracy

Hesitancy and refusal to be vaccinated were quantitatively related to the failure of health workers to provide appropriate and necessary information and support for vaccination (Table 1): “*I am hesitant to be vaccinated because I have not yet received relevant information about the vaccine*” (Male, hesitant). In addition, the communication offered by the health staff, but also the example given, seems to have a strong influence on the behaviour to be vaccinated:

- *“We need medical personnel to communicate with the population. They could have confidence if, for example, a doctor who is a specialist in his or her field communicates about the vaccine*. (Female, hesitant).
- *“The state should communicate widely through doctors and not through politicians. The state should communicate widely through doctors and not through politicians. Health personnel should be vaccinated and the population should be encouraged to do the same. For me, this is the best thing to do*. (Female, refusal).

This communication by health professionals is very important because it will reassure the population for a better success of the vaccination campaigns: “The day I am reassured, I will vaccinate myself” (Female, hesitant).

The quality of information was also highlighted in the qualitative interviews, and the conspiracy theories were prominent: “*The day before yesterday, I was shown someone who works in this field explaining that the vaccines sent to Africa are not the best and that they carry undesirable risks*” (Male, refusal). Some even went so far as to question whether the vaccine received by the state authorities is the same as the one that the population will use: “*There is no proof that the vaccine received by the health authorities and the state authorities are the same vaccines used for the population. For me, it is only a decoy because we saw on Whatsapp, a shot used in Germany that shows a person vaccinated with a fake syringe containing nothing and making people believe that the person was vaccinated”* (Male, hesitant).

#### Perceived effectiveness

Respondents thought that possible variants could complicate vaccination: “*The vaccine may be risky because it has been said that the Covid-19 virus can mutate. But how can you find your vaccine so quickly? If we find the cure and the virus takes another form too*…*I am not convinced of the effectiveness of this vaccine*.*”* (Female, refusal)

*“They won’t be as effective as they should be because the vaccine was designed for the variant that was here first. But if another variant comes along, this vaccine won’t be able to do anything about it because the virus has changed*” (Female, hesitant)

*On the* other hand, some people have reservations about certain vaccines: “From *what people are saying about the Chinese vaccine and the Russian vaccine, they have a lot more confidence in those two vaccines than the others. I don’t know if that’s the reality, but that’s from what I hear people say on both sides*.” (Male, hesitant).

## Discussion

The international public health and economic impact of COVID-19 has prompted private and governmental organizations to work together to address the pandemic. Significant investments have been made in developing vaccines against COVID-19 [29]. Nevertheless, hesitation in addition of accessibility about the COVID-19 vaccine may limit global efforts to control the pandemic and its adverse health and socioeconomic effects. [14]. Our study showed that 12.9% of individuals hesitate to be vaccinated, and 32.8% would not take the vaccine when it became available in Senegal. These results are similar to those of a study conducted in New Zealand and those of a systematic review and meta-analysis including 13 countries [16,30]. However, compared to these studies conducted in the USA, Portugal, and Great Britain, the proportion of refusals is higher in our study [4,16,30–32]. This result is even more worrying as the systematic review and meta-analysis of Robinson et al. [16] showed that the percentages of vaccine refusals and hesitation increased as the pandemic progressed. This situation could be confirmed by the disposal of thousands of doses of expired COVID-19 vaccine in October 2021 because the number of people vaccinated is quite small [18].

Vaccine hesitancy was associated with female gender in our study in Senegal. This result is similar to those found in New Zealand, Israel, China, the UK, the USA, Qatar, and Portugal. [3,4,30,32–34]. This disparity could be explained by the fact that women perceive a lower risk of the disease [3]. In addition, several reports and medias have shown higher risks of complications, infectiousness, and death from COVID-19 in men [35]. Therefore, women may be less likely to be affected by this disease. In addition, the finding in this study that women are more likely to show reluctance to be vaccinated is of concern as women play a central role in the vaccination of children.

Hesitancy and refusal to be vaccinated were related to living in large cities (Dakar, Thiès, and Diourbel). These same perceptions were noted in our previous study, which showed that the more regions are affected by the pandemic, the less confidence respondents have in the government and the less effective they perceive the measures to be [22]. As of December 2, 2021, these three most populated regions of Senegal will account for more than 80% of the cases of COVID-19 in the entire country [18]. One might have thought that vaccination intentions would be greater in these areas because of the burden of the pandemic. However, these results could be explained by the belief in a certain natural immunity, by a greater exposure to misinformation encountered on social networks, or by their higher standard of living than elsewhere in the country. The results of the first national seroprevalence survey in November 2021 may help us understand this perception.

The study showed that a bad attitude (thinking that it was not desirable and important to be vaccinated) towards vaccination was linked to hesitating and refusing vaccination. This poor attitude was mainly explained in the qualitative survey by rumours circulating—particularly on social networks—about the vaccine and the length of time it took to manufacture it. These reasons were consistent with the findings of several studies [3,17,36]. One report showed that the main topics of conversation related to vaccine hesitancy on Facebook and Twitter included posts about “dropouts,” people not showing up for their second injection, and parents resisting vaccinating their children “because of the low risk of COVID infection in their home.” [37]. Not surprisingly, there is a growing focus on the role of the media and in particular social media in shaping public opinion around COVID-19 and the vaccine. Social media, with its instant communication and access to a large audience, when combined with the ability to express oneself anonymously, offers immense potential for the spread of unverified and uncontrolled information. [34]. Public health organizations, health professionals, and media platforms should collaborate to ensure the accuracy of information, provide programs to improve health literacy levels to enable the target population to make an informed decision. Furthermore, the impact of these actions implies that strategies to overcome hesitancy can be framed in models that take into account these multi-faceted and multi-level factors [3].

The fact that individuals thought that vaccination would not help protect them from the virus was associated with reluctance and refusal to be vaccinated. Also, individuals who thought that the coronavirus vaccine might put their health at risk were more likely to refuse the vaccine. Indeed, several studies show that concerns about vaccine safety and efficacy appear to be important in vaccine intention [3,4,12,17,32,34]. This concern transcends socio-demographic aspects and countries. This concern led to the fact that some respondents to the survey wanted to “wait and see” whether vaccination was safe before getting vaccinated. Thus, effective communication about safety and efficacy, and greater transparency about vaccine development and distribution, including financial aspects, should be the cornerstone of all other strategies to ensure equitable mass immunization programs related to COVID-19 [38–40].

The hesitancy and refusal to vaccinate was also related to the fact that individuals thought that most people important to them would not think they should be vaccinated against COVID-19, and that health workers do not provide the appropriate and necessary information and support for vaccination. Several studies have examined the role of these factors consistent with our study [40–49]. Health professionals appeared to be a reliable source of information. Their recommendations [42,46,47,49] and support from family and friends [43,47] play an important role in influencing their perceptions and behaviours towards vaccination. These results suggest that health professionals (especially general practitioners and paediatricians) need to be better involved in vaccination campaigns to support people and help them make informed decisions.

This study is not without its limitations. It only involved people with mobile phones, thus excluding the most marginalized populations. In addition, the cross-sectional nature of the data limits our ability to draw conclusions about causal links. However, the sample is representative of the Senegalese population and the use of mixed methods allowed for a better understanding of the results and the organization of the arguments.

## Conclusion

The results of the study show that the factors related to hesitation and refusal to be vaccinated against COVID-19 in Senegal are diverse and complex. Reducing them will help to ensure better vaccination coverage as antigens become available. Therefore, governments and health authorities should intensify their efforts to encourage vaccine confidence and reduce misinformation. However, continued monitoring of COVID-19 vaccine hesitancy and refusal in the coming months will be critical to adjusting measures to address vaccine hesitancy, thereby ensuring adequate uptake of the vaccine. Finally, the current situation in Africa shows that there are still many challenges to vaccine accessibility for which the international community must act urgently. In addition to acceptance, accessibility is the second phase of the vaccine coverage coin that should not be forgotten.

## Data Availability

All data produced in the present study are available upon reasonable request to the authors

## Authors’ contributions

VR, AF and AJ designed the study. IG, AID, BK and MFB participated in the data collection. MFB and BK analyzed the data. MFB, BK, AF and VR interpreted the data. MFB wrote the article. All authors critically reviewed the article for intellectual content. All authors gave final approval to the submitted version.

## Financing

This research was conducted within the framework of the ARIACOV programme (Support for the African Response to the Covid-19 Epidemic), financed by the French Development Agency through the “COVID-19 - Health in Common” initiative.

## Conflict of Interest Statement

The authors declare that they have no known financial conflicts of interest or personal relationships that might appear to influence the work reported in this article.

## Ethical approval

The study received approval from the National Health Research Ethics Committee of Senegal (SEN/20/23).

### Appendices

**Appendix 1:** Socio-demographic characteristics of individuals in the qualitative survey (N=30)

| Features | Hesitant | Refusal |
| --- | --- | --- |
| **Sex** |  |  |
| Male | 9 | 9 |
| Female | 6 | 6 |
| **Age** |  |  |
| < 25 years | 3 | 3 |
| 25-59 years old | 10 | 10 |
| >=60 years | 2 | 2 |
| **Education** |  |  |
| Without instruction | 6 | 6 |
| Primary | 3 | 3 |
| Secondary | 4 | 4 |
| Superior | 2 | 2 |
| **Region** |  |  |
| Dakar | 5 | 5 |
| Outside Dakar | 10 | 10 |
| **Total** | **15** | **15** |

**Appendix 2:** Distribution of individuals by characteristics (N=607)

|  | n (%) |
| --- | --- |
| **Age:** |  |
| Under 25 years old | 140 (23.1) |
| 25-59 years old | 407 (67.1) |
| 60 years and over | 60 (9.9) |
| **Sex:** |  |
| Female | 241 (39.7) |
| Male | 366 (60.3) |
| **Quintile:** |  |
| Poorer | 81 (13.3) |
| Poor | 78 (12.9) |
| Medium | 131 (21.6) |
| Rich | 161 (26.5) |
| Richer | 156 (25.7) |
| **Region:** |  |
| Other regions | 257 (42.3) |
| Dakar-Diourbel-Thiès | 350 (57.7) |
| **Education:** |  |
| Without instruction | 253 (41.7) |
| Primary | 122 (20.1) |
| Secondary | 153 (25.2) |
| Superior | 79 (13.0) |
| **Have you ever received a vaccine as an adult?** |  |
| Yes | 173 (28.5) |
| No | 434 (71.5) |
| **I think it is important to get vaccinated:** |  |
| Yes | 444 (73.1) |
| No | 163 (26.9) |
| **I think it is useful to get the vaccine to protect against COVID-19:** |  |
| Yes | 377 (62.1) |
| No | 230 (37.9) |
| **I think it's responsible to get vaccinated against COVID-19:** |  |
| Yes | 510 (84.0) |
| No | 97 (16.0) |
| **I believe that the future COVID-19 vaccine will not pose a health risk:** |  |
| Yes | 203 (33.4) |
| No | 404 (66.6) |
| **I think it is advisable to get the COVID-19 vaccine:** |  |
| Yes | 408 (67.4) |
| No | 197 (32.6) |
| **Intention to be vaccinated:** |  |
| Yes | 330 (54.4) |
| Hesitates | 78 (12.9) |
| Refusal | 199 (32.8) |
| **Getting the COVID-19 vaccine will help protect me from the virus:** |  |
| Yes | 357 (58.8) |
| No | 250 (41.2) |
| **Getting vaccinated will help fight the spread of the coronavirus:** |  |
| Yes | 358 (59.0) |
| No | 249 (41.0) |
| **Getting the COVID-19 vaccine will help protect my loved ones from the virus:** |  |
| Yes | 349 (57.5) |
| No | 258 (42.5) |
| **I think the people who are going to create the COVID-19 vaccine are going to make sure it's safe:** |  |
| Yes | 330 (54.4) |
| No | 277 (45.6) |
| **The coronavirus vaccine could put my health at risk:** |  |
| Yes | 134 (22.1) |
| No | 473 (77.9) |
| **The coronavirus vaccine may have side effects:** |  |
| Yes | 226 (37.2) |
| No | 381 (62.8) |
| **When the vaccine is offered, most of the people important to me (family, friends) would think I need to get the** |  |
| **COVID-19 vaccine:** |  |
| Yes | 324 (53.4) |
| No | 283 (46.6) |
| **I think it will be easy for me to access the health care provider to get the coronavirus vaccine if I want it:** |  |
| Yes | 470 (77.4) |
| No | 137 (22.6) |
| **How much would you trust the health care providers who would give you a COVID-19 vaccine? Would you say you trust them:** |  |
| Yes | 441 (72.7) |
| No | 166 (27.3) |
| **Over the next few months, I will be learning about the COVID-19 vaccine on a regular basis:** |  |
| Yes | 521 (85.8) |
| No | 86 (14.2) |
| **Have you seen or heard anything bad about COVID-19 vaccines?** |  |
| Yes | 348 (57.4) |
| No | 258 (42.6) |
| **Health workers will need to provide appropriate and necessary information and support for immunization:** |  |
| Yes | 586 (96.5) |
| No | 21 (3.5) |
| **Free for all Senegalese:** |  |
| No | 90 (14.8) |
| Yes | 517 (85.2) |
| **Do you have confidence in the government of Senegal to fight the coronavirus epidemic?** |  |
| Yes | 170 (28.0) |
| No | 437 (72.0) |

**Appendix 3:**
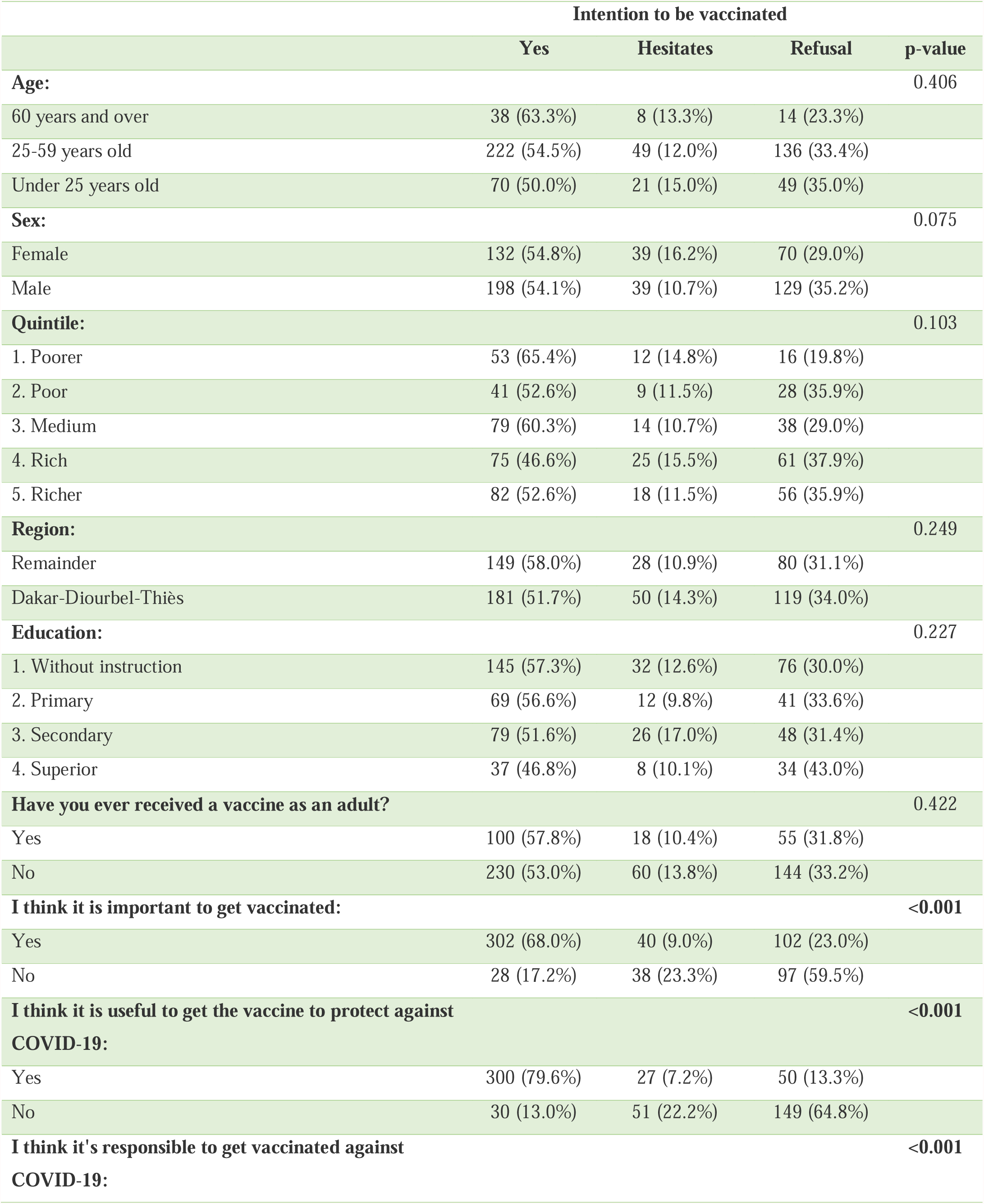
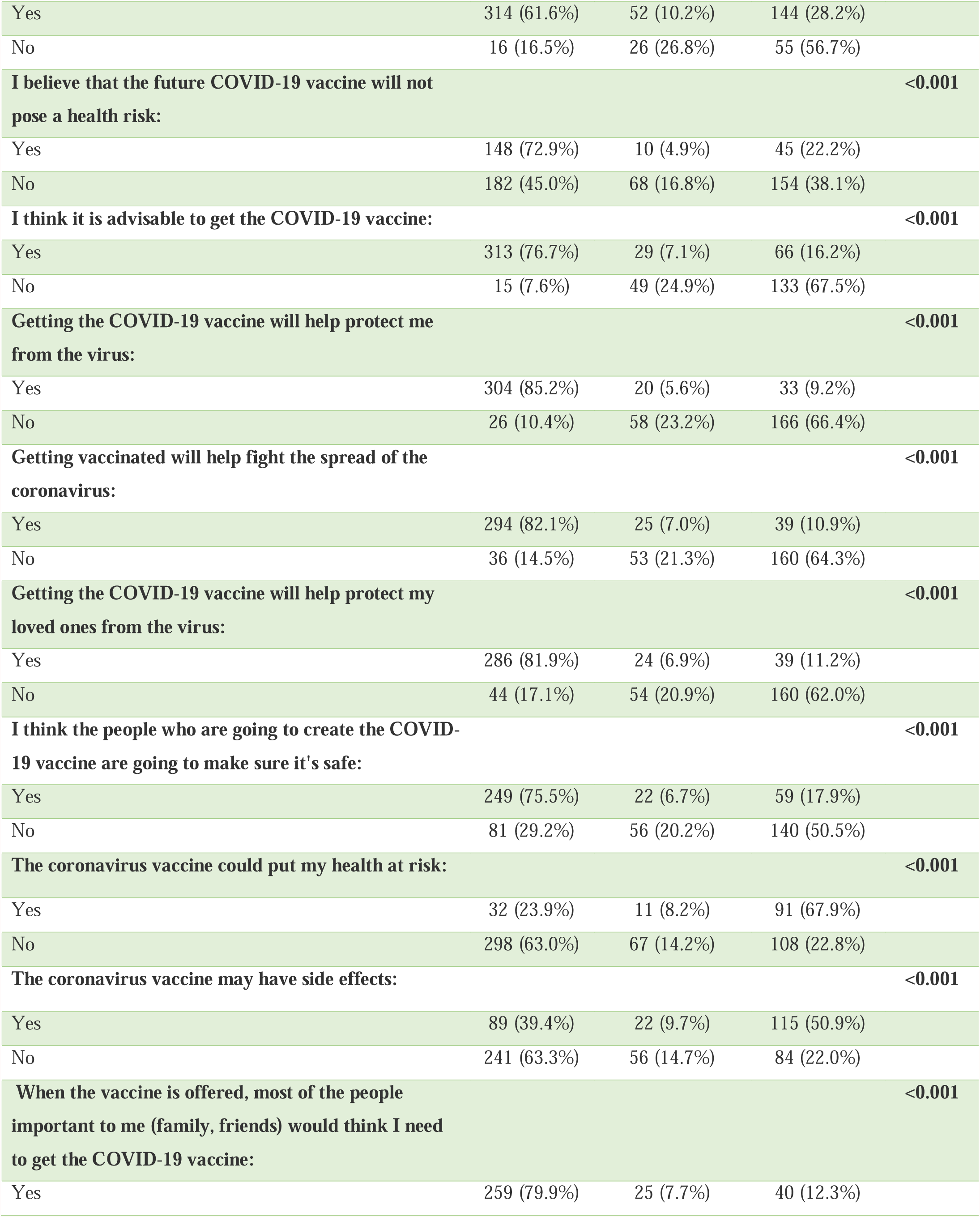
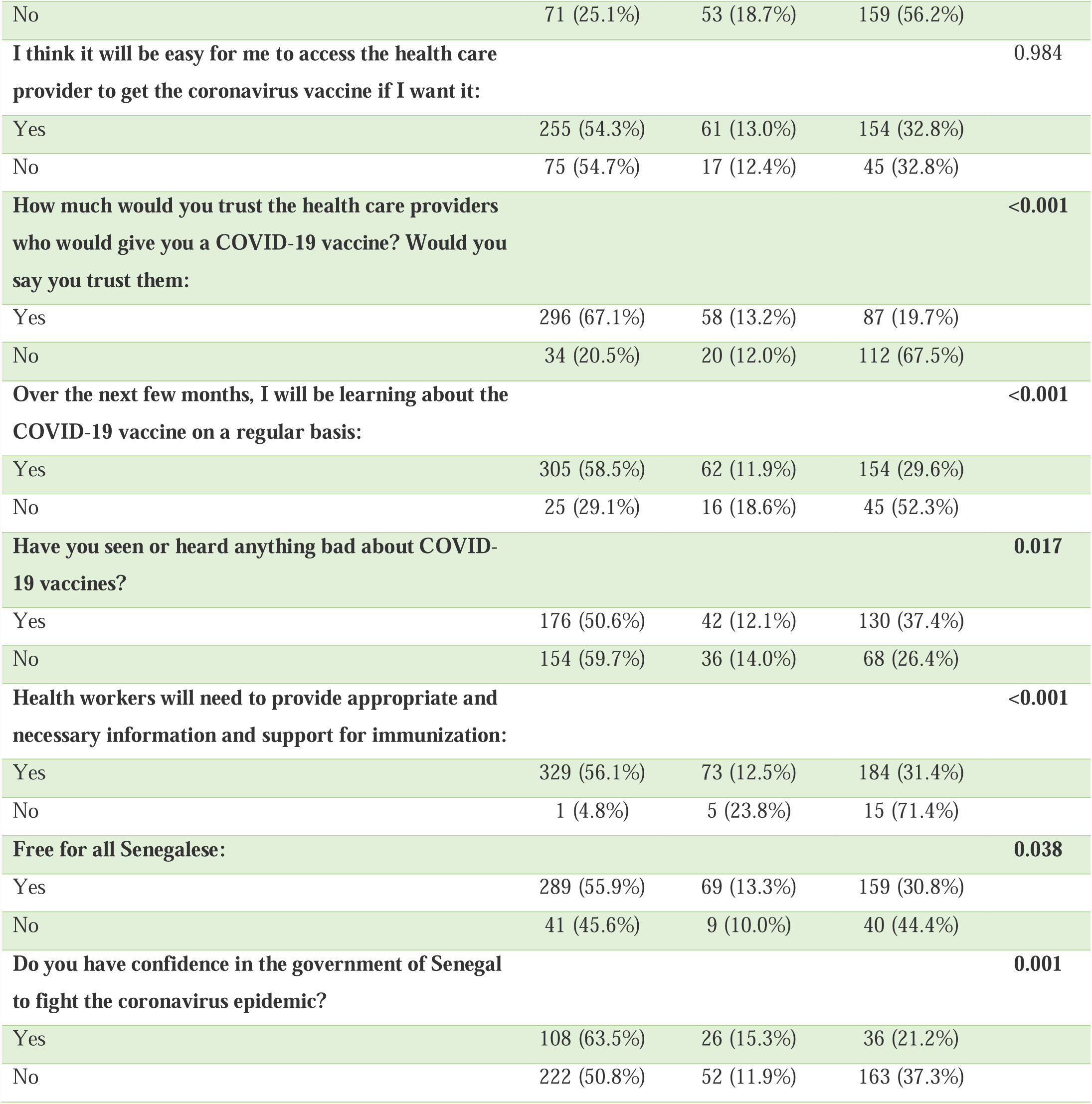
Distribution of individuals by characteristics and intention to be vaccinated (N=607)

## Notes

### Competing Interest Statement

The authors have declared no competing interest.

